# An Analysis for Key Indicators of Reproducibility in Radiology

**DOI:** 10.1101/19005074

**Authors:** Bryan D. Wright, Nam Vo, Johnny Nolan, Austin L. Johnson, Tyler Braaten, Daniel Tritz, Matt Vassar

## Abstract

**Background:** Given the central role of radiology in patient care, it is important that radiological research is grounded in reproducible science. It remains unexamined whether there is a lack of reproducibility or transparency in radiologic research.

**Purpose:** The purpose of this study was to analyze published radiology literature for the presence or absence of key indicators of reproducibility.

**Methods:** This cross-sectional, retrospective study was performed by conducting a search of the National Library of Medicine to identify publications contained within journals in the field of Radiology. Journals that were not written in English or MEDLINE indexed were excluded from the analysis. Studies published from January 1, 2014 to December 31, 2018 were used to generate a random list of 300 publications for this meta-analysis. A pilot-tested, Google form was used to evaluate key indicators of reproducibility in the queried publications.

**Results:** Our initial search returned 295,543 records, from which 300 were randomly selected for analysis. Of these 300 records, 294 met the inclusion criteria. Among the empirical publications, 5.6% contained a data availability statement (11/195, 95% CI: 3.0-8.3), 0.51% provided clearly documented raw data (1/195), 12.0% provided a materials availability statement (23/191, 8.4-15.7), none provided analysis scripts, 4.1% provided a preregistration statement (8/195, 1.9-6.3), 2.1% provided a protocol statement (4/195, 0.4-3.7), and 3.6% were preregistered (7/195, 1.5-5.7).

**Conclusion:** Our findings demonstrate that key indicators of reproducibility are missing in the field of radiology. Thus, the ability to reproduce radiological studies may be problematic and may have potential clinical implications.

## Introduction

The field of radiology plays a significant role in the diagnosis, monitoring, and treatment of numerous disease processes. The importance of radiology to the field of medicine is evident by the large annual expenditures on imaging, estimated to be 10% of total healthcare costs in the United States.(1) Advancements in imaging modalities and diagnostic testing are predicated upon robust and trustworthy research. Yet, the field of radiology has been known for low-level-evidence study designs, with randomized trials, multicenter studies, and meta-analyses making up the smallest portion of publications (0.8% to 1.5%).(2) With movement toward patient-centered, evidence-based care, efforts are needed to ensure the robustness and reproducibility of radiology research.

Reproducibility – defined as the ability to conduct an independent replication study and reach the same or similar conclusions as the study in question(3,4) – gained national attention after the majority of 1,500 surveyed scientists reported an inability to reproduce another scientist’s experiment and half being unable to reproduce their own experiments.(5) In radiology research, a lack of reproducibility has been partly attributed to imaging datasets that are too small to power significant findings, models lacking independent validation, and improper separation of training and validation data.(6) Such practices may go undetected by editors, peer reviewers, and readers and contribute to downstream effects, such as irreproducible results and perpetuated errors in subsequent studies.

Given the central role of radiology in patient care, reproducible radiology research is necessary. In this study, we investigated radiology publications for key factors of reproducibility and transparency. Findings may be used to evaluate the current climate of reproducible research practices in the field and contribute baseline data for future comparison studies.

## Materials and Methods

Our investigation was designed as a cross-sectional, meta-research study to evaluate specific indicators of reproducibility and transparency in radiology. The study methodology is a replication of work done by Hardwick et al.,(7) with minor adjustments. Our analysis did not utilize human subjects, thus this investigation was not subject to institutional review board oversight.(8) Guidelines detailed by Murad and Wang were used for the reporting of our meta-research.(9) The Preferred Reporting for Systematic Reviews and Meta-Analyses (PRISMA) guidelines were used as necessary.(10) We supplied all protocols, raw data, and pertinent materials on the Open Science Framework (https://osf.io/n4yh5/).

### Journal and Study Selection

One investigator (DT) used the subject term tag “Radiology[ST]” to search the National Library of Medicine (NLM) catalog on June 5, 2019. To meet the inclusion criteria, journals had to be MEDLINE indexed and written in the English language. DT extracted the electronic ISSN number (or linking ISSN if the electronic version was unavailable) for the included journals. PubMed was searched using the list of ISSN (PubMed contains the MEDLINE collection) on June 5, 2019 to identify publications. Publications from January 1, 2014 to December 31, 2018 were included. A random sample of 300 were selected to have data extracted with additional publications available as needed (https://osf.io/4hq87/). With the goal of creating a diverse spectrum of publications for our study, restrictions were not placed on specific study types.

### Training

Three investigators (BW, NV, and JN) underwent rigorous, in-person training led by DT on data extraction and study methodology to ensure reliability between investigators. Training included a review of the following: objectives of the study, design of the study, protocol, Google form used for data extraction, and the process of extracting data. The process for extracting data was demonstrated via the use of two example publications. All investigators who underwent training independently conducted a blinded, duplicate extraction of data from two further example publications. Once the mock data extraction was completed, the investigators (BW, NV, and JN) convened and resolved any discrepancies. The entire training session was recorded from the presenters point of view and was posted online for investigators to reference (https://osf.io/tf7nw/).

### Data Extraction

Once all required training was completed, data was extracted from publications. Data extraction began on June 9, 2019 and was completed on June 20, 2019. One investigator (BW) performed data extraction on 300 publications with the other two investigators (NV and JN) extracting from 150 each. We divided the publications into two categories: (1) publications with empirical data (e.g., clinical trial, cohort, case series, case reports, case-control, secondary analysis, chart review, commentary [with data analysis], and cross-sectional) and (2) publications without empirical data (e.g., editorials, commentaries [without reanalysis], simulations, news, reviews, and poems). For the sake of this study, imaging protocols with no patients or intervention were considered nonempirical. Different study designs resulted in variation in the data collected from individual publications. We analyzed nonempirical studies for the following characteristics: funding source(s), conflict of interest declarations, open access, and journal impact factor. Case reports and case series are not typically expected to be reproducible with a prespecified protocol.(11) As a result, data were extracted from them in an identical manner as publications that lacked empirical data. There was no expectation for meta-analyses and systematic reviews to contain additional materials, and so, the materials availability indicator was excluded from their analysis. For the purpose of our study, data were synonymous with raw data and considered unaltered data directly collected from an instrument. Investigators were prompted by the data extraction form to identify the presence or absence of necessary prespecified indicators of reproducibility, which is available here: https://osf.io/3nfa5/. The Google form used in this study included additional options beyond those contained in the form used by Hardwick et al.(7) Our form had additional study designs, such as case series, cohort, secondary analysis, meta-analysis/systematic review, chart review, and cross-sectional. Sources of funding were more specific to include non-profit, public, hospital, university, and private/industry. Following data extraction, all three investigators convened and resolved any discrepancies by consensus. Though unneeded, a third party was readily available for adjudication.

### Assessing Open Access

We systematically evaluated the accessibility of a full-text version of each publication. The Open Access Button (https://openaccessbutton.org/) was used to perform a search using publication title, DOI, and/or PubMed ID. If this search failed to provide an article, then Google Scholar and PubMed were searched using these parameters. If an investigator was still unable to locate a full-text publication, it was deemed inaccessible.

### Evaluation of Replication and Citation in Research Synthesis

Web of Science was searched for all studies containing empirical data. Once located on the Web of Science, we searched for the following: (1) the number of times a publication was used as part of a subsequent replication study and (2) the number of times a publication was cited in a systematic review/meta-analysis. Titles, abstracts and full-text manuscripts available on the Web of Science were used to analyze if a study was cited in a systematic review/meta-analysis or a replication study.

### Statistical Analysis

Statistics from each category of our analysis were calculated using Microsoft Excel. Excel functions were used to provide quantitative analysis with results characterized by frequencies, percentages, and 95% confidence intervals.

## Results

### Journal and Publication Selection

The NLM catalog search identified 144 radiology journals, but only 64 met the inclusion criteria. Our PubMed search for included publications included in these journals identified 295,543 radiology publications with 53,328 being published within the time frame. We randomly sampled 300, but 6 publications were inaccessible, even with institutional access. Of the eligible publications, 215 contained empirical data, and 79 did not (Figure 1). Publications without empirical data were excluded from certain analyses because they could not be assessed for reproducibility characteristics. Furthermore, 20 publications were identified as either case studies or case series; these research designs are unable to be replicated and were excluded from the analysis of study characteristics. Study reproducibility characteristics were analyzed for the remaining 195 radiology publications (Table 1).

**Table 1:** Reproducibility Indicators. The indicators measured for the articles varied depending on its study type. More details about extraction and coding are available here: https://osf.io/x24n3/

| *Reproducibility Indicator* |  | *Role in producing transparent and reproducible science.* |
| --- | --- | --- |
| **Articles** |  |  |
| Article accessibility:<br>Articles were assessed for open accessibility, paywall access, or unable to access full text | All (n=300) | Ease of access to publications enables interdisciplinary research by removing access barriers. Full text access allows for validation through reproduction. |
| **Funding** |  |  |
| Funding statement:<br>Presence of funding sources of the study | All included publications (n=294) | Funding provides researchers the ability to create new experiments and tangibly investigate their ideas. However, funding sources can play a role in influencing how researchers conduct and report their study (e.g. scientific bias), which necessitates its disclosure. |
| **Conflict of Interest (COI)** |  |  |
| COI statement:<br>Presence of conflict of interest statement | All included publications (n=294) | Conflict of interest conveys the authors' potential associations that may affect the experimental design, methods, and analyses of the outcomes. Thus, full disclosure of possible conflicts allows for unbiased presentation of their study. |
| **Data** |  |  |
| Data statement:<br>Presence of a data availability statement, retrieval method, comprehensibility, and content | Empirical publications‡ (n=195) | Raw data availability facilitates independent verification of research publications. It can improve accountability of outcomes reported and integrity of the research published. |
| **Materials** |  |  |
| Materials statement:<br>Presence of a materials availability statement, retrieval method, and accessibility | Empirical publications ¶ (n=191) | Materials are the tools used to conduct the study. Lack of materials specification impedes the ability to reproduce a study. |
| **Pre-registration** |  |  |
| Pre-registration statement:<br>Presence of statement indicating registration, retrieval method, accessibility, and contents (hypothesis, methods, analysis plan) | Empirical publications‡ (n=195) | Pre-registration explicitly reports aspects of the study design prior to the commencement of the research. Pre-registration functions as a way to limit selective reporting of results and prevents publication biases and P-hacking. |
| **Protocols** |  |  |
| Protocol statement:<br>Assessed for statement indicating protocol availability, and if available, what aspects of the study are available (hypothesis, methods, analysis plan) | Empirical publications‡ (n=195) | Reproducibility of a study is dependent on the accessibility of the protocol. A protocol is a highly detailed document that contains all aspects of the experimental design which provides a step by step guide in conducting the study. |
| **Analysis scripts** |  |  |
| Analysis scripts statement:<br>Presence of analysis script availability statement, retrieval method, and accessibility | Empirical publications‡ (n=195) | Analysis scripts are used to analyze data obtained in a study through software programs such as R, Python, and Matlab. Analysis scripts provide step by step instructions to reproduce statistical results. |
| **Replication** |  |  |
| Replication statement:<br>Presence of statement indication a replication study. | Empirical studies‡ (n = 195) | Replication studies provide validation to previously done publications by determining whether similar outcomes can be acquired. |
| † Excludes publications that have no empirical data.<br>‡ Excludes case studies and case series<br>¶ Excludes meta-analyses and systematic reviews, which materials may not be relevant, in addition to ‡. |  |  |

**Figure 1:**
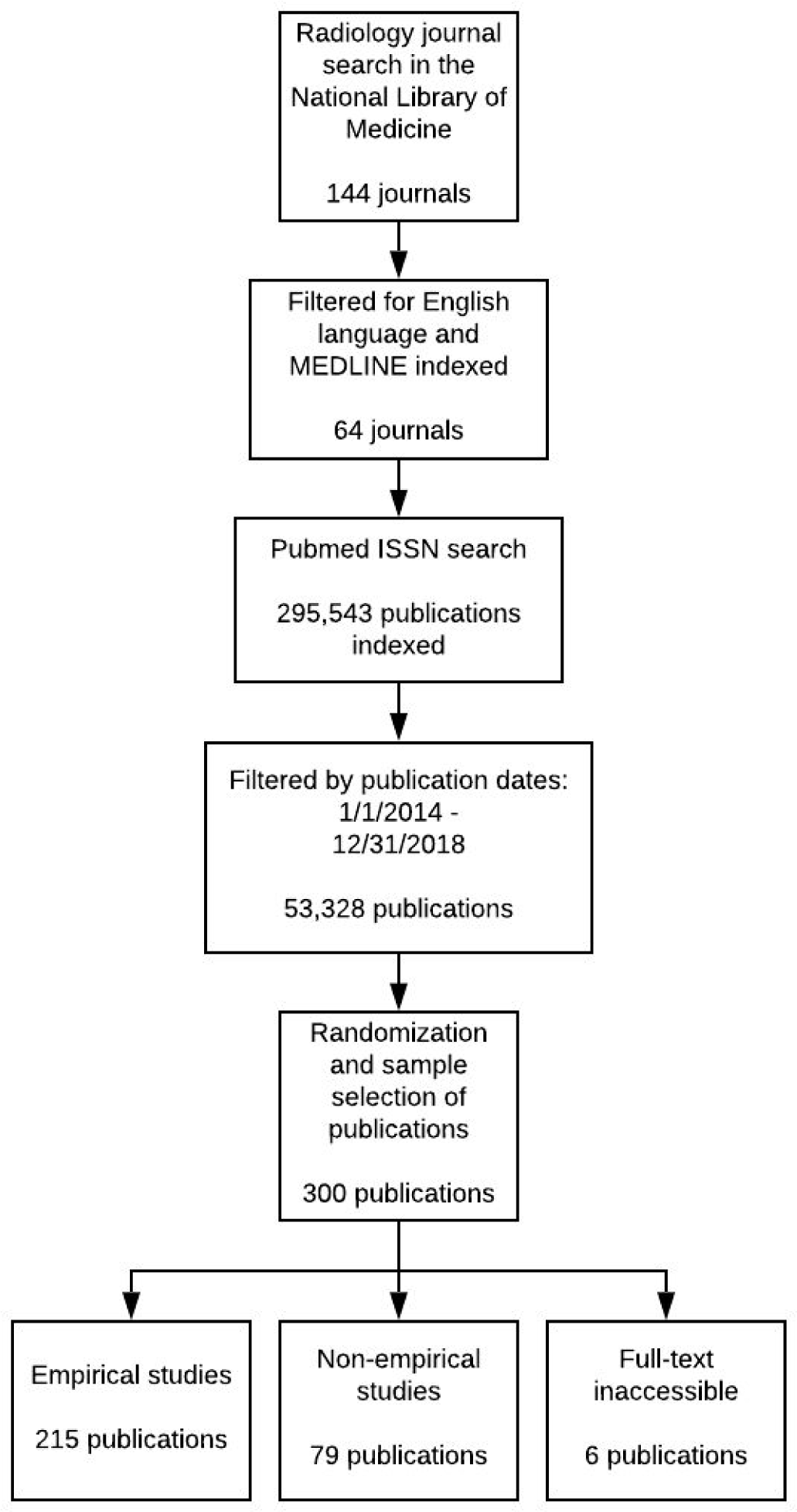
Publication selection process.

### Sample Characteristics

From our sample of 294 radiology publications, the publishing journals had a median 5-year impact factor of 2.824 (interquartile range: 1.765-3.718). Study designs of sampled publications are presented in Table 2. Most publications originated from the United States (209/294, 71.1%), followed by the United Kingdom (44/294, 15.0%). The majority of authors were from the United States (102/294, 34.7%), followed by China (19/294, 6.5%). Humans were the most common test subjects (167/294, 56.8%).

**Table 2:** Characteristics of Included Publications

| **Characteristic** |  | **N (%)** |
| --- | --- | --- |
| **Type of Study N=294** | No empirical data | 79 (26.9) |
|  | Clinical trial | 55 (18.7) |
|  | Laboratory | 44 (15.0) |
|  | Chart Review | 42 (14.3) |
|  | Cohort | 19 (6.5) |
|  | Case study | 18 (6.1) |
|  | Survey | 12 (4.1) |
|  | Cost-effectiveness/decision analysis | 7 (2.4) |
|  | Case control | 6 (2.0) |
|  | Cross-sectional | 5 (1.7) |
|  | Meta-analysis | 4 (1.4) |
|  | Case series | 2 (0.7) |
|  | Multiple | 1 (0.3) |
|  | Other | 0 (0.0) |
| **Test Subjects N=294** | Humans | 167 (56.8) |
|  | Neither | 116 (39.5) |
|  | Animals | 11 (3.7) |
|  | Both | 0 (0) |
| **Country of journal publication N=294** | US | 209 (71.1) |
|  | UK | 44 (15.0) |
|  | Germany | 6 (2.0) |
|  | France | 5 (1.7) |
|  | Japan | 3 (1.0) |
|  | Canada | 3 (1.0) |
|  | Italy | 2 (0.7) |
|  | India | 1 (0.3) |
|  | Other | 21 (7.14) |
| **Country of corresponding author N=294** | US | 102 (34.7) |
|  | China | 19 (6.5) |
|  | Germany | 17 (5.8) |
|  | Japan | 17 (5.8) |
|  | Australia | 15 (5.1) |
|  | South Korea | 14 (4.8) |
|  | Turkey | 13 (4.4) |
|  | Canada | 13 (4.4) |
|  | UK | 9 (3.1) |
|  | Netherlands | 9 (3.1) |
|  | France | 8 (2.7) |
|  | Switzerland | 7 (2.4) |
|  | India | 5 (1.7) |
|  | Italy | 5 (1.7) |
|  | Spain | 3 (1.0) |
|  | Unclear | 2 (0.7) |
|  | Other | 36 (12.2) |
| **Open Access N=300** | Yes found via Open Access Button | 80 (26.7) |
|  | Yes found article via other means | 71 (23.7) |
|  | Could not access through paywall | 149 (49.7) |
| **5 Year Impact Factor N=272** | Median | 2.824 |
|  | 1st quartile | 1.765 |
|  | 3rd quartile | 3.718 |
|  | Interquartile range | 1.953 |
| **Most Recent Impact Factor Year N=300** | 2017 | 266 |
|  | 2018 | 10 |
|  | Not found | 24 |
| **Most Recent Impact Factor**<br>**N=276** | Median | 2.758 |
|  | 1st quartile | 1.823 |
|  | 3rd quartile | 3.393 |
|  | Interquartile range | 1.57 |
| **Cited by Systematic Review or**<br>**Meta-Analysis N=211** | No Citations | 194 (91.9) |
|  | A Single Citation | 17 (8.1) |
|  | One to Five Citations | 8 (3.8) |
|  | Greater than Five Citations | 0 (0) |
|  | Excluded in SR or MA | 0 (0) |
| **Cited by Replication Study**<br>**N=211** | No Citations | 211 (100) |
|  | A Single Citation | 0 (0) |
|  | One to Five Citations | 0 (0) |
|  | Greater than Five Citations | 0 (0) |
|  | Excluded in SR or MA | 0 (0) |

**Table 3:** Reproducibility-related Characteristics of Included Publications

| Characteristics |  | N (%) | 95% CI |
| --- | --- | --- | --- |
| **Open Access N=300** | Paywall access required | 149 (49.7) | 44.0-55.3 |
|  | Open access button | 80 (26.7) | 21.7-31.7 |
|  | Found via other means | 71 (23.7) | 18.9-28.5 |
| **Funding N=294** | No funding statement | 176 (59.9) | 54.3-65.4 |
|  | Public | 47 (16.0) | 11.8-20.1 |
|  | No funding received | 28 (9.5) | 6.2-12.8 |
|  | Multiple funding sources | 22 (7.5) | 4.5-10.5 |
|  | Non-profit | 7 (2.4) | 0.7-4.1 |
|  | University | 6 (2.0) | 0.4-3.6 |
|  | Private/Industry | 6 (2.0) | 0.4-3.6 |
|  | Hospital | 2 (0.7) | 0.0-1.6 |
| **Conflict of Interest statement N=294** | No statement | 100 (34.0) | 28.7-39.4 |
|  | No conflicts | 156 (53.1) | 47.4-58.7 |
|  | Conflicts | 38 (12.9) | 9.1-16.7 |
| **Data Availability N=195** | No statement | 184 (94.4) | 91.7-97.0 |
|  | Available* | 11 (5.6) | 3.0-8.3 |
|  | Not available | 0 (0.0) | 0 |
| **Materials Availability N=191** | No Statement | 168 (88.0) | 84.3-91.6 |
|  | Available* | 23 (12.0) | 8.4-15.7 |
|  | Not available | 0 (0) | 0-0 |
| **Pre-registration N=195** | No statement | 187 (95.9) | 93.7-98.1 |
|  | Available* | 7 (3.6) | 1.5-5.7 |
|  | Not available | 1 (0.5) | 0-1.3 |
| **Protocol Availability**<br>**N=195** | Not available | 191 (97.9) | 96.3-99.6 |
|  | Available* | 4 (2.1) | 0.4-3.7 |
| **Analysis Scripts N=195** | No statement | 195 (100) | 100 |
|  | Not available | 0 | 0 |
|  | Available | 0 | 0 |
| **Replication Studies**<br>**N=195** | Novel study | 195 (100) | 100 |
|  | Replication | 0 (0) | 0 |
| *Reproducibility-related characteristics that are available contain further specifications within Table 4.<br>Abbreviations: CI, Confidence Interval |  |  |  |

### Reproducibility Related Characteristics

Nearly half of the eligible publications required paywall access (149/300, 49.7%). Open access publications were found via the Open Access Button 26.7% of the time (80/300) or via Google Scholar or Pubmed 23.7% of the time (71/300). More than half of the publications failed to provide a funding statement (176/294, 59.9%). Public funding accounted for 16% of analyzed publications (47/294). Authors reported having no conflicts of interest (COI) in the majority of publications (No COI: 156/294, 53.1% v. COI: 38/294, 12.9%). No COI statement was provided 34.0% of the time (100/294). Data availability was reported in 11 publications (11/195, 5.6%), but only nine had accessible data (9/11, 81.8%). Complete, raw data was located in 0.51% of empirical publications (1/195). A materials availability statement was included in 23 publications (23/191, 12.0%), but only 18 provided access to materials used in the study (18/23, 78.3%). Most publications did not include a preregistration statement (8/195, 4.1%) or protocol statement (4/195, 2.1%). Specifications of reproducibility-related characteristics are reported in Table 4. Among the 195 publications containing empirical data, none provided analysis scripts for reproducing statistical results (0/195, 0%). None of the publications reported being a replication (0/195, 0%). Most publications were not cited in systematic review or meta-analysis (194/211, 91.9%), whereas a few publications were cited in either a single publication (17/211, 8.1%) or between one and five publications (8/211, 3.8%). None of the publications were cited in replication studies (0/211, 0%).

**Table 4:** Specifications of Reproducibility-related Characteristics of Included Publications

| Characteristic-Specifications |  |  | N (%) |
| --- | --- | --- | --- |
| **Data Available (n=11)** | Data retrieval | Supplementary information hosted by the journal | 10 (90.9) |
|  |  | Online third party | 1 (9.1) |
|  |  | Upon request from authors | 0 (0.0) |
|  | Data accessibility | Yes, data could be accessed and downloaded | 9 (81.8) |
|  |  | No, data count not be accessed and downloaded | 2 (18.2) |
|  | Data documentation§ | Yes, data files were clearly documented | 8 (88.9) |
|  |  | No, data count not be accessed and downloaded | 1 (11.1) |
|  | Raw data availability§ | No, data files do not contain all raw data | 6 (66.7) |
|  |  | Unclear if all raw data was available | 2 (22.2) |
|  |  | Yes, data files contain all raw data | 1 (11.1) |
| **Materials available (n = 23)** | Materials retrieval | Supplementary information hosted by the journal | 15 (65.2) |
|  |  | Online third party | 4 (17.4) |
|  |  | In the paper | 4 (17.4) |
|  | Materials accessibility | Yes, materials could be accessed and downloaded | 18 (78.3) |
|  |  | No, materials not be accessed and downloaded | 5 (21.7) |
| Protocol available<br>(n=4) | Protocol contents | Methods | 4 (100.0) |
|  |  | Analysis plan | 0 (0.0) |
|  |  | Hypothesis | 0 (0.0) |
| Pre-registration<br>available (n=7) | Registry | Clinicaltrials.gov | 6 (85.7) |
|  |  | Brazilian Registry of<br>Clinical Trials | 1 (14.3) |
|  | Pre-registration<br>accessibility | Yes, pre-registration<br>could be accessed | 6 (85.7) |
|  |  | No, pre-registration<br>could not be accessed | 1 (14.3) |
|  | Pre-registration<br>contents \|\| | Methods | 6 (100.0) |
|  |  | Hypothesis | 2 (33.3) |
|  |  | Analysis Plan | 0 (0.0) |
| §Data specifications for documentation and raw data availability are provided if data was made accessible. |  |  |  |
| \|\| Pre-registration specifications for contents are provided if pre-registration was made accessible. |  |  |  |

## Discussion

Our cross-sectional investigation found that key transparency and reproducibility-related factors were rare or entirely absent among our sample of publications in the field of radiology. No analyzed publications reported an analysis script, a minority provided access to materials, few were preregistered, and only one provided raw data. While concerning, our findings are similar to those found in the social science and biomedical literature.(7,12,13) Here, we discuss two of the main findings from our study.

One factor that is important for reproducible research is the availability of all raw data. In radiology, clinical data and research data, when shared, are often stored and processed in online repositories. Picture archives and communication systems allow researchers to observe details of image acquisition, patient positioning, image depth, and bit depth of an image; but proprietary formats, large file sizes, and protected health information can complicate sharing of radiology data.(14) However, picture archiving and communication systems and software prototypes that combine the benefits of clinical and research designs have been developed for the purpose of storing, retrieving and sharing functional imaging data.(15) Additionally, the rarity of data sharing may also be due to an absence of specific recommendations from radiology journals, with a survey of 18 general imaging journals finding only one with policies requesting data at submission.(16) By improving data sharing in radiology, others have the ability to critically assess the trustworthiness of data analysis and the interpretation of results.(17) With more than 30 radiology and imaging journals being listed as International Committee of Medical Journal Editors (ICMJE) members, the ICMJE could have a substantial influence by enforcing their data sharing policy and commending the dissemination of research results and datasets.(18–21) A recent survey by the European Society of Radiology research committee found that 98% of respondents would be interested in sharing data, yet only 23 institutions (34%) had previously shared clinical trial data. From the data shared by these 23 institutions, at least 44 additional original works have been published.(22)

A second factor lacking in radiology literature was access to detailed analysis scripts. In radiology research, many analytic decisions exist regarding data management, radiomic identification, biomarker identification with validation, and sharing necessary items (data, coding, statistics, and protocol).(23–26) Radiomic study findings cannot be accurately reproduced or validated without an exact description of the steps taken to measure and analyze the data points. A systematic review of 41 radiomic studies found that 16 failed to report detailed software information, two failed to provide image acquisition settings, and eight lacked detailed descriptions about any preprocessing modifications. These three methodological areas are important in radiological imaging studies as they can alter results significantly, thus decreasing the reproducibility of study findings.(27) A recent study by Carp et al. further tested possible variations in radiology image analysis by using a combination of five preprocessing and five modeling decisions for data acquisition in functional magnetic resonance imaging. This modification in data collection created almost 7,000 unique analytical pathways with varying results.(28) For research findings to be reproducible, a detailed analysis script with explicit software information and methodological decision making is necessary, this could be accomplished through public repositories and “containers” that replicate the study calculations in real time and are applicable to other data sets.(29) Investigators should be encouraged to take notes of analysis coding and scripts so as to be able to create a detailed explanation to be published with the study.(30–32) Providing such analysis scripts could improve the reproducibility of study outcomes and serve as a guide for future radiology publications to follow.(26)

### Implications Moving Forward

Our sample indicates that there is room for improvement in reporting reproducibility-related factors in radiology research. Ninety percent of scientists agree that science is currently experiencing a “reproducibility crisis,” with “more robust experimental designs and better statistics” seen as possible solutions.(5) To improve reporting of experimental designs, we encourage authors to follow available reporting guidelines.(33)(34,35) In radiology, reliability and agreement studies are prevalent as interobserver agreement between radiologists is measured to identify the potential for errors in treatments or diagnostic imaging.(36) The Guidelines for Reporting Reliability and Agreement Studies (GRRAS) is a 15-item checklist required for study findings to be accurately interpreted and reproduced in reliability and agreement studies(37). Items such as sample selection, study design, and statistical analysis are often omitted by authors.(36,38,39) Researchers have had success with using the GRRAS, specifically, but reporting guidelines, in general, provide the framework for studies to be understood by readers, reproduced by researchers, used by doctors, and included in systematic reviews.(38,40,41)

### Better Statistics

The current reproducibility crisis is, in part, tied to poor statistics. In psychology, a large-scale reproducibility study found that only 39 of 100 original psychology studies could be successfully replicated.(42) In response, the Association for Psychological Science has pioneered several innovations, such as statistical verification programs, and statistical advisors, to provide expertise on manuscripts with sophisticated statistics or methodological techniques, and to promote reproducibility within psychology.(43) Based on our findings, it is possible that radiology may be experiencing similar transparency and reproducibility problems and should consider promoting improved statistical practices by using a statistician to assist in the review process. StatReviewer – an automated review of statistical tests and appropriate reporting – may aid peer-reviewers who are not formally trained in detecting relevant statistical errors or detailed methodological errors.(44)

## Strengths and Limitations

Regarding the strengths of this study, we randomly sampled a large selection of radiology journals. Our double data extraction methodology was performed in similar fashion to systematic reviews by following the *Cochrane Handbook*.(48) Complete raw data and all relevant study materials are provided online to ensure transparency and reproducibility. Regarding its limitations, our analysis included only 300 of the 53,328 returned publications in the radiology literature; thus, our results may not be generalizable to publications in time periods outside of our search or other medical specialties. Our study focused on analyzing the transparency and reproducibility of the published literature in radiology, and as such, we relied solely on information reported within the publications. Therefore, it cannot be assumed that reproducibility-related factors are not available upon request from the author. Had we contacted the corresponding authors of the 300 analyzed publications, it is plausible we could have obtained more information.

## Conclusion

With the potential lack of transparency and reproducibility practices in radiology, opportunities exist to improve radiology research. Our results indicate important factors for reproducibility and transparency are frequently missing in radiology publications, leaving room for improvement. Methods to improve reproducibility and transparency are practical and applicable to many research designs.

## Data Availability

We supplied all protocols, raw data, and pertinent materials on the Open Science Framework (https://osf.io/n4yh5/).

## References

1. Howell W. Imaging utilization trends and reimbursement. Diagn Imaging. 2014;

2. Rosenkrantz AB, Pinnamaneni N, Babb JS, Doshi AM. Most Common Publication Types in Radiology Journals:: What is the Level of Evidence? Acad Radiol. 2016;23(5):628–633.

3. Pitcher RD. The Role of Radiology in Global Health. In: Mollura DJ, Culp MP, Lungren MP, editors. Radiology in Global Health: Strategies, Implementation, and Applications. Cham: Springer International Publishing; 2019. p. 157–174.

4. WHO | Medical imaging. World Health Organization; 2017; https://www.who.int/diagnostic_imaging/en/. Accessed June 27, 2019.

5. Baker M. 1,500 scientists lift the lid on reproducibility. Nature. 2016;533(7604):452–454.

6. Aerts HJWL. Data Science in Radiology: A Path Forward. Clin Cancer Res. 2018;24(3):532–534.

7. Hardwicke TE, Wallach JD, Kidwell M, Ioannidis J. An empirical assessment of transparency and reproducibility-related research practices in the social sciences (2014-2017). 2019.http://dx.doi.org/10.31222/osf.io/6uhg5.

8. Electronic Code of Federal Regulations - US Department of Health and Human Services’ Code of Federal Regulation 45 CFR 46.102(d). https://www.ecfr.gov/cgi-bin/retrieveECFR?gp=&SID=83cd09e1c0f5c6937cd9d7513160fc3f&pitd=20180719&n=pt45.1.46&r=PART&ty=HTML#se45.1.46_1102 in effect July 19, 2018.

9. Murad MH, Wang Z. Guidelines for reporting meta-epidemiological methodology research. Evid Based Med. 2017;22(4):139–142.

10. Liberati A, Altman DG, Tetzlaff J, et al. The PRISMA statement for reporting systematic reviews and meta-analyses of studies that evaluate health care interventions: explanation and elaboration. J Clin Epidemiol. 2009;62(10):e1–e34.

11. Wallach JD, Boyack KW, Ioannidis JPA. Reproducible research practices, transparency, and open access data in the biomedical literature, 2015–2017. PLoS Biol. Public Library of Science; 2018;16(11):e2006930.

12. Iqbal SA, Wallach JD, Khoury MJ, Schully SD, Ioannidis JPA. Reproducible Research Practices and Transparency across the Biomedical Literature. PLoS Biol. 2016;14(1):e1002333.

13. Wallach JD, Boyack KW, Ioannidis JPA. Reproducible research practices, transparency, and open access data in the biomedical literature, 2015–2017. PLoS Biol. Public Library of Science; 2018;16(11):e2006930.

14. Read Metadata from DICOM Files - MATLAB & Simulink. https://www.mathworks.com/help/images/read-metadata-from-dicom-files.html. Accessed August 4, 2019.

15. Doran SJ, d’Arcy J, Collins DJ, et al. Informatics in radiology: development of a research PACS for analysis of functional imaging data in clinical research and clinical trials. Radiographics. 2012;32(7):2135–2150.

16. Sardanelli F, Alì M, Hunink MG, Houssami N, Sconfienza LM, Di Leo G. To share or not to share? Expected pros and cons of data sharing in radiological research. Eur Radiol. 2018;28(6):2328–2335.

17. Warren E. Strengthening Research through Data Sharing. N Engl J Med. 2016;375(5):401–403.

18. Naudet F, Sakarovitch C, Janiaud P, et al. Data sharing and reanalysis of randomized controlled trials in leading biomedical journals with a full data sharing policy: survey of studies published inThe BMJandPLOS Medicine. BMJ. 2018. p. k400http://dx.doi.org/10.1136/bmj.k400.

19. Federer LM, Belter CW, Joubert DJ, et al. Data sharing in PLOS ONE: An analysis of Data Availability Statements. PLoS One. 2018;13(5):e0194768.

20. 0000-0003-1953-, 0000-0002-7378-. Making Progress Toward Open Data: Reflections on Data Sharing at PLOS ONE | EveryONE: The PLOS ONE blog. EveryONE. 2017. https://blogs.plos.org/everyone/2017/05/08/making-progress-toward-open-data/. Accessed June 20, 2019.

21. ICMJE | Journals stating that they follow the ICMJE Recommendations. http://www.icmje.org/journals-following-the-icmje-recommendations/. Accessed August 12, 2019.

22. Bosserdt M, Hamm B, Dewey M. Clinical trials in radiology and data sharing: results from a survey of the European Society of Radiology (ESR) research committee. Eur Radiol. 2019;http://dx.doi.org/10.1007/s00330-019-06105-y.

23. Piccolo SR, Frampton MB. Tools and techniques for computational reproducibility. Gigascience. 2016;5(1):30.

24. Garijo D, Kinnings S, Xie L, et al. Quantifying reproducibility in computational biology: the case of the tuberculosis drugome. PLoS One. 2013;8(11):e80278.

25. Gronenschild EHBM, Habets P, Jacobs HIL, et al. The effects of FreeSurfer version, workstation type, and Macintosh operating system version on anatomical volume and cortical thickness measurements. PLoS One. 2012;7(6):e38234.

26. Parmar C, Barry JD, Hosny A, Quackenbush J, Aerts HJWL. Data Analysis Strategies in Medical Imaging. Clin Cancer Res. 2018;24(15):3492–3499.

27. Traverso A, Wee L, Dekker A, Gillies R. Repeatability and Reproducibility of Radiomic Features: A Systematic Review. Int J Radiat Oncol Biol Phys. 2018;102(4):1143–1158.

28. Carp J. On the plurality of (methodological) worlds: estimating the analytic flexibility of FMRI experiments. Front Neurosci. 2012;6:149.

29. Poldrack RA, Gorgolewski KJ, Varoquaux G. Computational and Informatic Advances for Reproducible Data Analysis in Neuroimaging. Annu Rev Biomed Data Sci. Annual Reviews; 2019;2(1):119–138.

30. Triphan S, Biederer J, Burmester K, et al. Raw data and analysis scripts for “Design and application of an MR reference phantom for multicentre lung imaging trials.” heiDATA; 2018.http://dx.doi.org/10.11588/DATA/FHOCRZ.

31. Triphan SMF, Biederer J, Burmester K, et al. Design and application of an MR reference phantom for multicentre lung imaging trials. PLoS One. 2018;13(7):e0199148.

32. Gorgolewski KJ, Poldrack RA. A Practical Guide for Improving Transparency and Reproducibility in Neuroimaging Research. PLoS Biol. 2016;14(7):e1002506.

33. The CARE Guidelines: Consensus-based Clinical Case Reporting Guideline Development | The EQUATOR Network. http://www.equator-network.org/reporting-guidelines/care/. Accessed August 13, 2019.

34. Reporting guidelines | The EQUATOR Network. http://www.equator-network.org/reporting-guidelines/. Accessed August 13, 2019.

35. Guidelines for reporting reliability and agreement studies (GRRAS) were proposed | The EQUATOR Network. https://www.equator-network.org/reporting-guidelines/guidelines-for-reporting-reliability-and-agreement-studies-grras-were-proposed/. Accessed August 13, 2019.

36. Kottner J, Audigé L, Brorson S, et al. Guidelines for Reporting Reliability and Agreement Studies (GRRAS) were proposed. J Clin Epidemiol. 2011;64(1):96–106.

37. Kottner J, Audige L, Brorson S, et al. Guidelines for Reporting Reliability and Agreement Studies (GRRAS) were proposed. International Journal of Nursing Studies. 2011. p. 661–671http://dx.doi.org/10.1016/j.ijnurstu.2011.01.016.

38. Gerke O, Möller S, Debrabant B, Halekoh U, Odense Agreement Working Group. Experience Applying the Guidelines for Reporting Reliability and Agreement Studies (GRRAS) Indicated Five Questions Should Be Addressed in the Planning Phase from a Statistical Point of View. Diagnostics. 2018. p. 69 http://dx.doi.org/10.3390/diagnostics8040069.

39. Cronin P, Rawson JV. Review of Research Reporting Guidelines for Radiology Researchers. Acad Radiol. 2016;23(5):537–558.

40. What is a reporting guideline? | The EQUATOR Network. http://www.equator-network.org/about-us/what-is-a-reporting-guideline/. Accessed August 13, 2019.

41. Oster NV, Carney PA, Allison KH, et al. Development of a diagnostic test set to assess agreement in breast pathology: practical application of the Guidelines for Reporting Reliability and Agreement Studies (GRRAS). BMC Women’s Health. 2013.http://dx.doi.org/10.1186/1472-6874-13-3.

42. Open Science Collaboration. Estimating the reproducibility of psychological science. Science. American Association for the Advancement of Science; 2015;349(6251):aac4716.

43. APS: Leading the Way in Replication and Open Science. Association for Psychological Science. https://www.psychologicalscience.org/publications/observer/obsonline/aps-reproducibility-and-replication-initiatives.html. Accessed June 30, 2019.

44. Stat Reviewer. http://www.statreviewer.com/. Accessed August 1, 2019.

45. Klein RA, Ratliff K, Vianello M, et al. Investigating variation in replicability: A “many labs” replication project. Open Science Framework. 2014.

46. Klein RA, Vianello M, Hasselman F, et al. Many Labs 2: Investigating Variation in Replicability Across Samples and Settings. Advances in Methods and Practices in Psychological Science. SAGE Publications Inc; 2018;1(4):443–490.

47. Munafò MR, Nosek BA, Bishop DVM, et al. A manifesto for reproducible science. Nature Human Behaviour. 2017.http://dx.doi.org/10.1038/s41562-016-0021.

48. Higgins JPT, Green S. Cochrane Handbook for Systematic Reviews of Interventions. John Wiley & Sons; 2011.

